# Feasibility and effectiveness of daily temperature screening to detect COVID-19 in a large public university setting

**DOI:** 10.1101/2021.03.22.21254140

**Authors:** Shelley N. Facente, Lauren A. Hunter, Laura J. Packel, Yi Li, Anna Harte, Guy Nicolette, Shana McDevitt, Maya Petersen, Arthur L. Reingold

**Author notes:** **Corresponding Author:** Shelley N. Facente, School of Public Health, Division of Epidemiology and Biostatistics, University of California, Berkeley, 2121 Berkeley Way # 5302, Berkeley, CA 94720.

## Abstract

**Background:** Many persons with active SARS-CoV-2 infection experience mild or no symptoms, presenting barriers to COVID-19 prevention. Regular temperature screening is nonetheless used in some settings, including University campuses, to reduce transmission potential. We evaluated the potential impact of this strategy using a prospective University-affiliated cohort.

**Methods:** Between June and August 2020, 2,912 participants were enrolled and tested for SARS-CoV-2 by PCR at least once (median: 3, range: 1-9). Participants reported temperature and symptoms daily via electronic survey using a previously owned or study-provided thermometer. We assessed feasibility and acceptability of daily temperature monitoring, calculated sensitivity and specificity of various fever-based strategies for restricting campus access to reduce transmission, and estimated the association between measured temperature and SARS-CoV-2 test positivity using a longitudinal binomial mixed model.

**Results:** Most participants (70.2%) did not initially have a thermometer for taking their temperature daily. Across 5481 total person months, the average daily completion rate of temperature values was 61.6% (IQR: 41.8%–86.2%). Sensitivity for SARS-CoV-2 ranged from 0% (95%CI 0–9.7%) to 40.5% (95%CI 25.6–56.7%) across all strategies for self-report of possible COVID-19 symptoms on day of specimen collection, with corresponding specificity of 99.9% (95%CI 99.8–100%) to 95.3% (95%CI 94.7–95.9%). An increase of 0.1°F in individual mean body temperature on the same day as specimen collection was associated with 1.11 increased odds of SARS-CoV-2 positivity (95%CI 1.06–1.17).

**Conclusions:** Daily temperature monitoring was feasible and acceptable; however, the majority of potentially infectious individuals were not detected by temperature monitoring, suggesting that temperature screening is insufficient as a primary means of detection to reduce transmission of SARS-CoV-2.

## BACKGROUND

SARS-CoV-2 is a novel coronavirus that causes severe coronavirus disease 2019, otherwise known as COVID-19.^1^ One of the most challenging features of the COVID-19 epidemic to date is considerable pre-symptomatic^2^ and asymptomatic transmission,^3,4^ currently estimated to comprise anywhere from 6% to 41% of infectious individuals;^5^ many other individuals may experience only mild symptoms. This presents barriers to epidemic control by impeding rapid isolation of cases, and makes it necessary to develop nuanced screening approaches that will both limit infectious exposure and be a useful trigger for SARS-CoV-2 testing. Accordingly, a strong desire to have students and employees return in person to school or work has driven workplaces, businesses, and colleges to look for methods to rapidly assess risk of infection and prevent entry for those who are possibly infectious to others.

One common strategy to prevent transmission has been temperature checks, which have increased in popularity as a non-invasive measure to rapidly screen individuals for elevated body temperature (i.e., fever). Temperature screening has been used during other global outbreaks, including the severe acute respiratory syndrome (SARS) outbreak in 2003, the H1N1 influenza epidemic in 2009,^6^ and recent major Ebola virus disease outbreaks in Sub-Saharan Africa in 2014 and 2018.^7^ However, multiple studies have found low sensitivity or specificity of temperature monitoring during these prior epidemics,^8,9^ even in cases where fever was a very common symptom among people with the disease in question.^6,10,11^ Large infrared fever screening systems or no-contact temperature screening at building entrances^12^ and hospital entryways^13^ and wearable devices to continuously monitor individual temperature^14^ have all been widely employed in an attempt to prevent the spread of SARS-CoV-2, yet limited evidence exists concerning the sensitivity and specificity of these approaches for the current pandemic,^15^ especially on a university campus. Of note, the United States Food and Drug Administration (FDA) released a statement in June 2020 noting that non-contact temperature assessment devices “are not effective if used as the only means of detecting a COVID-19 infection.”^16^

A number of college campuses are now implementing systems that require students, faculty, and staff to attest that they are free of symptoms, and/or are afebrile before coming to campus.^17-21^ These policies almost universally rely on dichotomous temperature cutoffs for fever, e.g. temperature ≥100.4°F, in alignment with the CDC guidelines.^22^ Such strategies have numerous pitfalls, including reliance on an outdated sense of “normal” body temperature,^23^ disregard of the effect of time of day^24^ and ambient temperature on body temperature,^25^ disregard of numerous studies that have found meaningful variation in normal body temperature between individuals,^26,27^ and incentives to not voluntarily disclose symptoms in order to preserve access to work spaces and therefore safeguard career and financial stability.^28^

Given the rapid increase in reliance on temperature-based strategies to restrict campus access for prevention of COVID-19 spread, we aimed to assess the feasibility, acceptability, and effectiveness of temperature monitoring and other fever-based strategies to prevent spread of SARS-CoV-2 in a longitudinal cohort of 2,916 university-affiliated students and employees, known as the Berkeley COVID-19 Safe Campus Initiative.

## METHODS

### Study Setting and Population

Any students, faculty, or staff (including essential workers) who were affiliated with the University of California, Berkeley and were living in Berkeley or the surrounding counties during the summer of 2020 were eligible to enroll in the Safe Campus cohort. Enrollment was completed on a rolling basis beginning on June 1^st^ and lasting until July 17^th^, at which time 2,916 participants had enrolled and provided a specimen for at least one PCR test. All participants with at least one valid PCR test result who reported not having tested positive for SARS-CoV-2 prior to enrollment were included in this analysis (n=2,900).

### Survey Measures and Temperature Assessment

Participants provided information about thermometer availability during a baseline survey, and were told that if they did not already own a thermometer they would be provided with one when they reported to University Health Services for baseline specimen collection. They were asked to report quantitative temperature and any symptoms potentially related to COVID-19 (including “feeling feverish”) on electronic daily surveys via a HIPAA-compliant version of Research Electronic Data Capture (REDCap),^29,30^ from the day after they completed the baseline survey through study close August 18^th^. Participants were prompted each morning to complete their daily survey via e-mail or text message, depending on their preference, and student participants were provided with a $50 incentive for completing their baseline specimen collection and 10 daily surveys in order to encourage habit formation.^31,32^ Due to rolling enrollment with a fixed end date, participants were enrolled for different lengths of time, ranging from 28 to 77 days between baseline specimen collection and August 18^th^ (median: 54 days, IQR: 43 – 64 days). Beginning August 1^st^ participants were sent a message requesting them to complete an endline survey that included questions about feasibility and acceptability of recording their temperature each day.

### PCR Testing

All participants were asked to come to University Health Services (UHS) on the UC Berkeley campus for an oral/nasal midturbinate swab for SARS-CoV-2 polymerase chain reaction (PCR) testing at the start of the study, and participants who were students or essential workers were also asked to return for an endline PCR test in early August. Participants were also offered PCR testing on-demand at any time during the study, and were specifically asked to come to UHS for a PCR test if they reported a temperature of ≥100.4°F, said they were “feeling feverish,” reported other specific symptoms (dry cough, coughing up mucus, unusual pain or pressure in the chest, difficulty breathing, shortness of breath, unexplained trouble thinking or concentrating, or loss of sense of taste or smell), or reported a specific potential exposure to COVID-19. Participants were PCR tested a minimum of once and a maximum of 9 times during the study, with a median of 3 tests per person. PCR tests were all conducted at UC Berkeley’s Innovative Genomics Institute.^33^

### Statistical Analysis

We assessed potential impact of temperature-based screening on transmission of SARS-CoV-2 through calculating sensitivity, specificity, positive predictive value (PPV), and negative predictive value (NPV) for a range of strategies for detecting SARS-CoV-2 infection, including temperature greater than a range of thresholds for “fever” (≥100.4°F, ≥98.7°F, ≥99.7°F) on the day of specimen collection, temperature ≥98.7°F on the day of specimen collection or on any day up to three days prior to collection, a qualitative assessment of “feeling feverish” as a symptom, and being “symptomatic,” which for purposes of this analysis included reporting dry cough, coughing up mucus, fever, sweats, chills, sore throat, difficulty breathing, wheezing, shortness of breath, loss of sense of taste, loss of sense of smell, or at least three symptoms from a list of 35 symptoms potentially associated with COVID-19 (see Supplemental Table 1). The results of PCR testing were used as the gold standard for determining “true” SARS-CoV-2 positivity or negativity in relation to these performance measures, and 95% confidence intervals (CIs) were calculated using the Clopper Pearson method^34^ for sensitivity and specificity and asymptotic standard logit intervals^35^ for the predictive values, using the *bdpv* package within R.^36^ Adjusted logit intervals were used to compute intervals in the case where the predictive value was zero. A Fisher’s Exact Test for independence was used to test the null hypothesis that there was no difference in SARS-CoV-2 infection for people who would have been prohibited entry to campus as a result of screening for fever using the strategy in question, and those who would be permitted entry to campus.

**Table 1.** Characteristics of study participants, by personal thermometer availability.

| Characteristic |  | Total participants | Needed a thermometer<br># (%) |  |
| --- | --- | --- | --- | --- |
| **Total** |  | **2,912** | **2,026** | **(69.6%)** |
| **Cohort** | Student | 2,174 | 1,646 | (75.7%) |
|  | Essential worker | 268 | 165 | (61.6%) |
|  | Faculty/staff | 470 | 215 | (45.7%) |
| **Age (years)** | <21 | 881 | 730 | (82.9%) |
|  | 22-29 | 1,085 | 787 | (72.5%) |
|  | 30-39 | 446 | 258 | (57.8%) |
|  | 40-49 | 195 | 103 | (52.8%) |
|  | 50+ | 279 | 148 | (53.0%) |
| **Gender identity** | Woman<sup>a</sup> | 1,634 | 1,101 | (67.4%) |
|  | Man<sup>b</sup> | 1,171 | 862 | (73.6%) |
|  | Non-binary | 50 | 37 | (74.0%) |
| **Race/Ethnicity<sup>c</sup>** | Black | 102 | 82 | (80.4%) |
|  | Latinx | 395 | 301 | (76.2%) |
|  | Asian/Pacific Islander | 825 | 637 | (77.2%) |
|  | Native American/Am. Indian | 38 | 26 | (68.4%) |
|  | White | 1,802 | 1,180 | (65.5%) |
|  | Other | 91 | 68 | (74.7%) |
| **PCR<sup>d</sup> results** | All negative | 2,852 | 1,980 | (69.4%) |
|  | At least one positive | 60 | 46 | (76.7%) |

<sup>a</sup> Category likely includes some participants who are transwomen
<sup>b</sup> Category likely includes some participants who are transmen
<sup>c</sup> Categories are not mutually exclusive
<sup>d</sup> PCR = Polymerase Chain Reaction

We used a longitudinal binomial mixed model with a random intercept for each participant to examine the association between individually mean-centered quantitative temperature and SARS-CoV-2 PCR result. This association was assessed using a simple model with no additional covariates, as well as a model controlling for mean-centered local ambient temperature on the day body temperature was recorded, and age of the participant.

### Ethics

This research was approved by the UC Berkeley Committee for the Protection of Human Subjects (CPHS), protocols #2020-06-13349 (essential workers), #2020-05-13261 (students), and #2020-04-13238 (faculty/staff).

## FINDINGS

### Feasibility of daily thermometer usage

At enrollment, 70.3% of participants did not have access to a thermometer for daily use at home and needed to be provided one by the study. This was particularly true for students (76.7%), participants under age 30 (77.2%), and Black (80.4%), Asian/Pacific Islander (77.4%) and Latinx (76.3%) participants (Table 1). The majority (70.5%) of participants who had their own thermometer had an oral thermometer, followed by 12.8% with a forehead thermometer, 7.8% with an ear thermometer, 4.1% with a no-touch thermometer, and the remainder with other types. All participants who received a thermometer through the study used an oral thermometer.

Participants who were supplied thermometers by the study had consistently lower mean temperatures (p <0.001) in comparison to participants using their own thermometers (Figure 1): those with their own thermometer had a mean reported temperature of 97.8°F over the course of the study (IQR: 97.4 – 98.2°F) and participants with a study-supplied thermometers had a mean reported temperature of 97.6°F (IQR: 97.1 – 97.9°F). These mean temperatures were deemed reasonable, given that participants were instructed to measure their temperature in the morning, when mean body temperatures have been found to generally be below the typically considered 98.6°F.^23^

**Figure 1.**
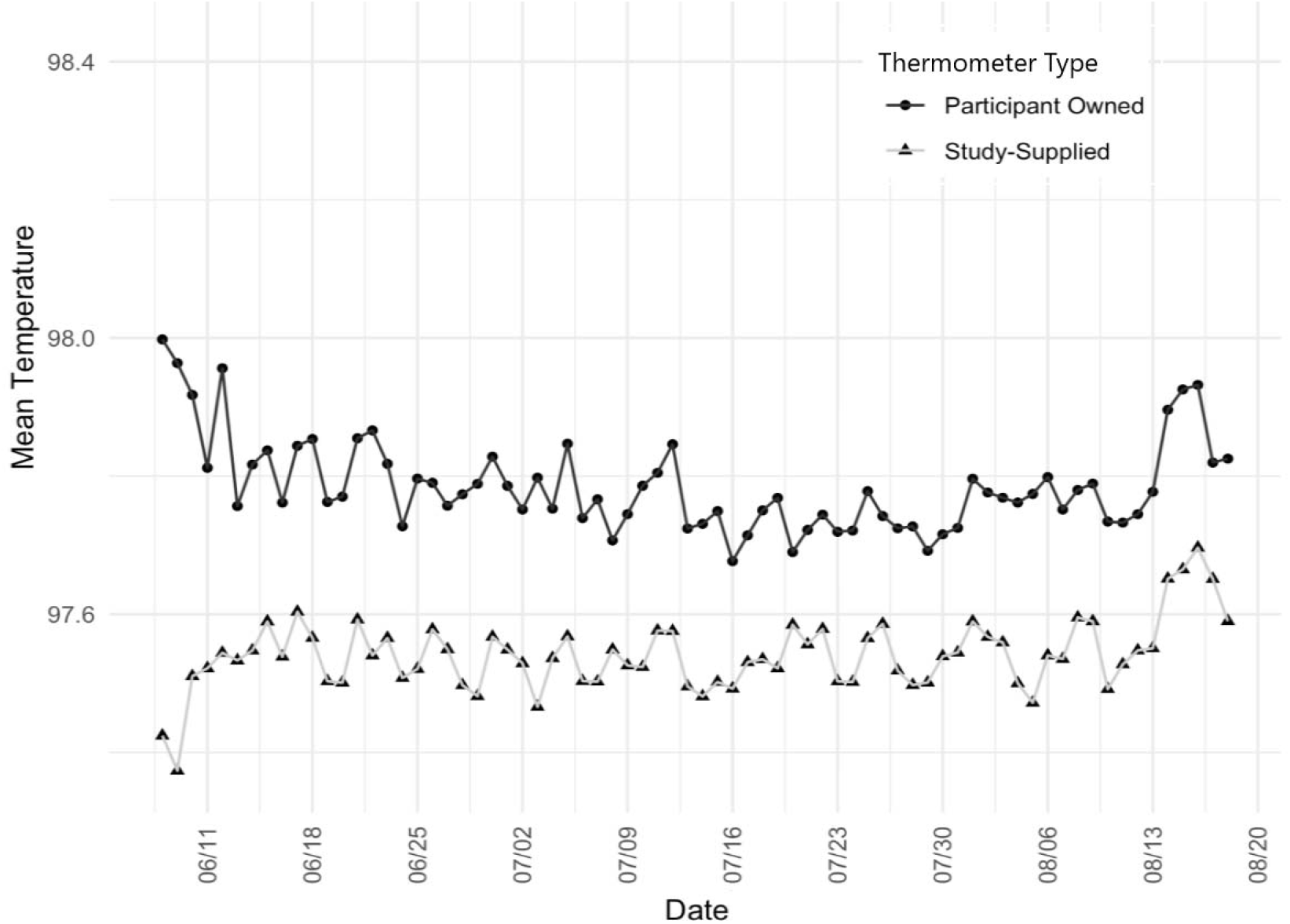
Mean body temperature per day, by type of thermometer

Participants reported their temperature measurement a median of 67.6% (IQR: 41.8 – 86.2%) of the total days they were enrolled (Figure 2, panel A), and more than 30% of participants (n = 885) recorded the temperature every day they completed at least some of the daily survey (Figure 2, panel B).

**Figure 2.**
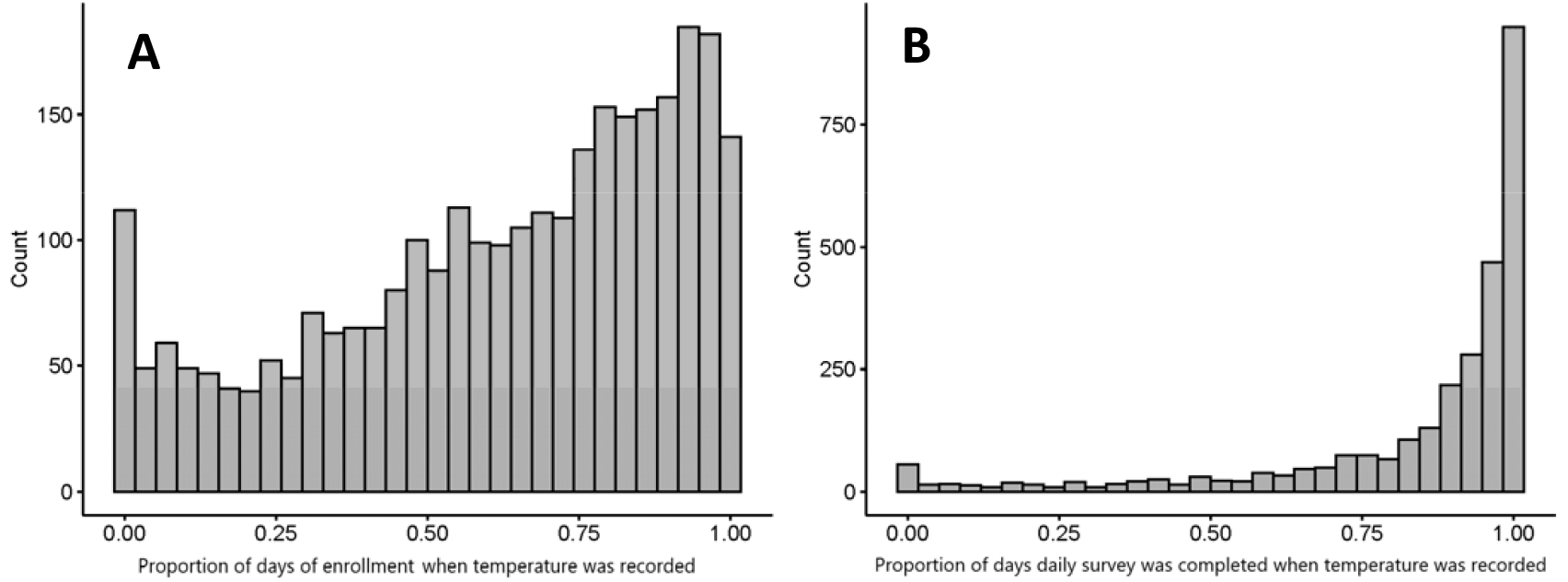
Participant count of the proportion of days quantitative temperature was recorded via the daily survey, per total days of enrollment (A) and total days for which the daily survey was at least partially completed (B).

### Acceptability of daily temperature monitoring

During the endline survey, participants were asked about the acceptability of daily temperature monitoring, and 92.9% reported it to be acceptable (n = 735) or totally acceptable (n = 1734). Only 35 participants out of 2,659 who took the endline survey (1.3%) found it to be unacceptable. When asked how likely they would be to continue to comply with daily temperature monitoring if UC Berkeley used this study as a model for campus-wide practice, 1079 people (40.5%) said they were “extremely likely” to continue, and 1027 (38.6%) said they would be “likely” to continue. Only 291 people (10.9%) said they were “unlikely” or “extremely unlikely” to comply with a request to continue monitoring their temperature daily. However, when asked about the “most difficult” aspect of their study participation at endline, daily temperature monitoring was the most frequently selected study component: 765 people (29.1%) chose daily temperature monitoring, compared to 14.2% who chose having oral/nasal swabs collected for PCR testing, the next most common answer.

### Effectiveness for detection of SARS-CoV-2 infection

More than a third (35%) of the 60 participants in our study who tested positive for SARS-CoV-2 did so at their baseline collection; as a result, positive test results were preceded by a median of six recorded temperatures (range 1-55, IQR 2-21). In comparison, negative tests were preceded by a median of 13 recorded temperatures (range 1-76, IQR 1-31)

Thirty-six (60.0%) of 60 positive tests for SARS-CoV-2 by PCR were preceded by a participant reported temperature the same day; 37 (61.7%) were preceded by at least one reported temperature in the three days prior to specimen collection.

Mean temperatures among participants testing positive ranged from 96.7°F -99.0°F during the study (Figure 3).Among the 56 participants who tested positive for SARS-CoV-2 and recorded their temperature at least once during the study period, only 4 participants measured at least one temperature above 100°F at any time, and a temperature ≥100°F was measured only 10 times out of 1,637 total measurements (0.6%) taken throughout the study among those testing positive. This is notable given the CDC-suggested threshold of 100.4°F.^22^

**Figure 3.**
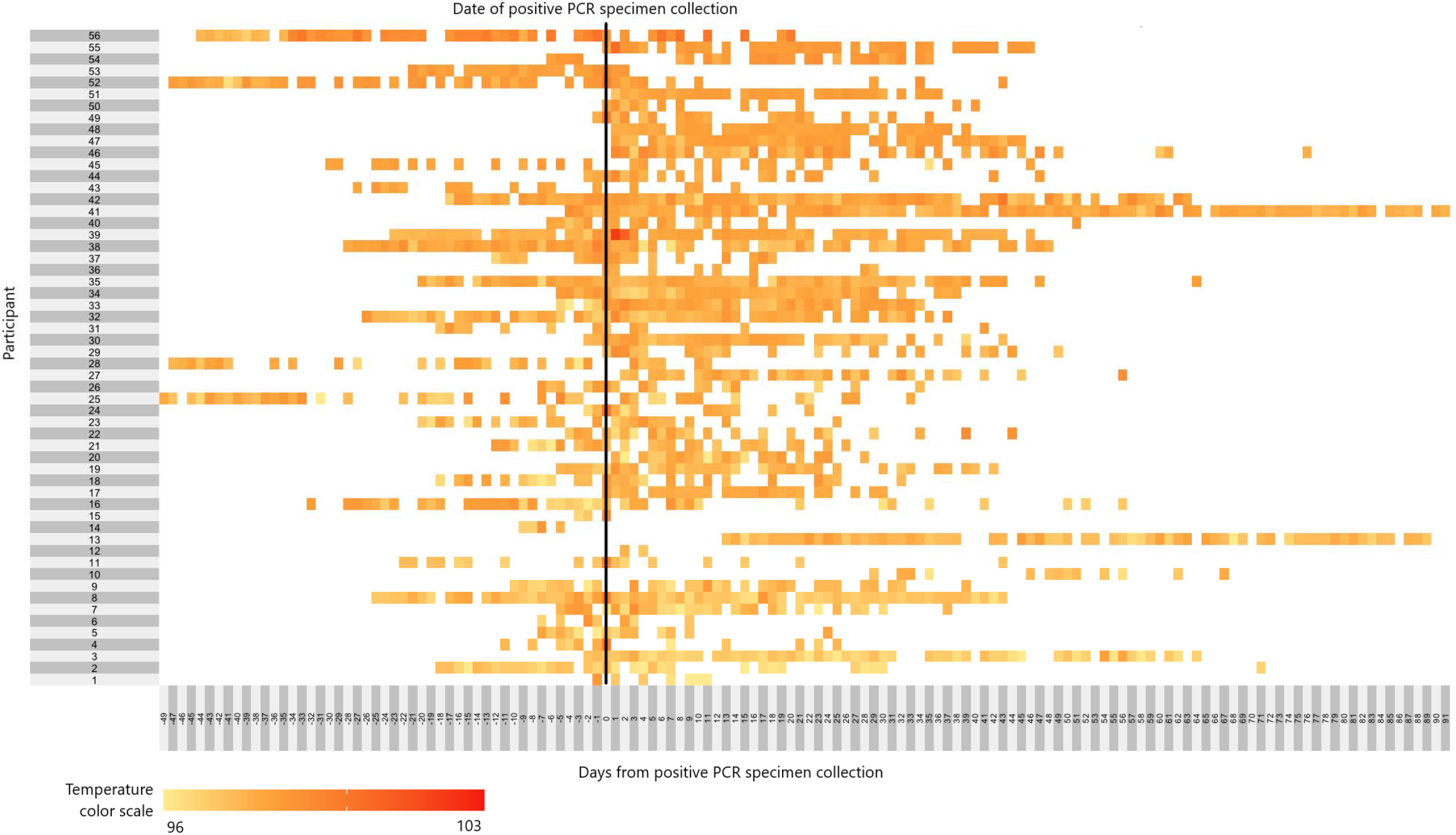
Heat map of temperatures measured each day before and after specimen collection at which 56* participants tested PCR positive for SARS-CoV-2. The vertical black line denotes the day that participant provided the specimen which would subsequently test positive for SARS-CoV-2. White boxes are days that the participant did not record a temperature (including days before they had enrolled in the study), and colored boxes range from yellow to red depending on the self-reported participant temperature, as found in the temperature color scale on the bottom left of the figure. * Note that 4 of the 60 participants who tested SARS-CoV-2 positive never recorded their body temperature through the study, and therefore have been omitted from this Figure

Sensitivity for detecting SARS-CoV-2 infection ranged from 0% (95% CI 0.0 – 9.7%) to 30.6% (95% CI 16.3 – 48.1%) for the strategies using various thresholds for fever on the same day as specimen collection, among the 4,330 people who recorded a temperature the same day as specimen collection (Table 2). Sensitivity predictably increased as the threshold for fever was lowered, with a resulting trade-off in specificity. Self-reported qualitative symptoms (“feeling feverish”) had a higher sensitivity (19.0%, 95% CI 8.6 – 34.1%) than either of the quantitative fever thresholds often used in practice (temperature ≥100.4°F or ≥99.7°F), and comparable specificity of 99.7% (95% CI 99.5 – 99.8%), compared to a specificity of 99.9% (95% CI 99.8 – 100%) for the ≥100.4°F threshold and 99.7% (95% CI 99.5 – 99.9%) for the ≥99.7°F threshold. Participants also reported information about qualitative fever symptoms the same day as their PCR specimen collection an additional 1,126 times, compared to quantitative temperature measurements. As would be expected, self-report of symptoms potentially indicating SARS-CoV-2 infection (including but not limited to fever) yielded the greatest sensitivity (40.5%, 95% CI 25.6 – 56.7%, with specificity of 95.3%, 95% CI 94.7 – 95.9%), but had a positive predictive value of only 6.3% (4.4 – 9.0%), well below that of self-reported “feeling feverish” alone (32.0%, 95% CI 17.8 – 50.9%).

**Table 2.** Performance characteristics of potential indicators of SARS-CoV-2 infection.

|  | Temperature<br>≥100.4°F<sup>a</sup> same day<br>as specimen<br>collection | Temperature<br>≥99.7°F<sup>b</sup> same day<br>as specimen<br>collection | Temperature<br>≥98.7°F<sup>c</sup> same day<br>as specimen<br>collection | Temperature<br>≥99.7°F same day<br>or any of 3 days<br>prior to specimen<br>collection | self-report fever<br>symptoms (i.e.<br>feeling feverish)<br>same day as<br>specimen collection | self-report<br>COVID-19 symptoms,<br>including fever<sup>d</sup><br>same day as<br>specimen collection |
| --- | --- | --- | --- | --- | --- | --- |
| Number of PCR<br>positive results with<br>relevant data recorded<br>(out of 60 total) | 36 | 36 | 36 | 37 | 42 | 42 |
| Number of PCR<br>negative results with<br>relevant data recorded<br>(out of 7612 total) | 4294 | 4294 | 4294 | 4487 | 5415 | 5415 |
|  | pt. est. (95% CI) | pt. est. (95% CI) | pt. est. (95% CI) | pt. est. (95% CI) | pt. est. (95% CI) | pt. est. (95% CI) |
| **Sensitivity** | 0.0% (0.0 - 9.7%) | 8.3% (1.8 - 22.5%) | 30.6% (16.3 - 48.1%) | 10.8% (3.0 - 25.4%) | 19.0% (8.6 - 34.1%) | 40.5% (25.6 - 56.7%) |
| **Specificity** | 99.9% (99.8 - 100%) | 99.7% (99.5 - 99.9%) | 93.0% (92.2 - 93.8%) | 99.4% (99.2 - 99.6%) | 99.7% (99.5 - 99.8%) | 95.3% (94.7 - 95.9%) |
| **PPV<sup>e</sup>** | 0.0% (0.0 - 59.4%) | 21.4% (7.4 - 48.6%) | 3.5% (2.2 - 5.8%) | 13.8% (5.6 - 30.6%) | 32.0% (17.8 - 50.9%) | 6.3% (4.4 - 9.0%) |
| **NPV<sup>f</sup>** | 99.2% (99.1 - 99.3%) | 99.2% (99.1 - 99.3%) | 99.4% (99.2 - 99.5%) | 99.3% (99.2 - 99.3%) | 99.4% (99.3 - 99.5%) | 99.5% (99.4 - 99.6%) |
| **Fisher's Exact test<sup>g</sup>**<br>**p-value** | -- | 0.0002 | <0.0001 | <0.0001 | <0.0001 | <0.0001 |

<sup>a</sup> This was the threshold used for triggering additional testing during the study
<sup>b</sup> This is one degree above what is conventionally considered "normal" body temp
<sup>c</sup> This is one degree above the mean temperature for our study participants over the full study period (mean = 97.6°F)
<sup>d</sup> For purposes of this analysis, participants were considered "symptomatic" if on their daily survey they reported dry cough, coughing up mucus, fever, sweats, chills, sore throat, difficulty breathing, wheezing, shortness of breath, loss of sense of taste, loss of sense of smell, or at least 3 symptoms from a list of 35 symptoms associated with COVID-19 and other illnesses (see Supplemental Table 1).
<sup>e</sup> PPV = Positive predictive value
<sup>f</sup> NPV = Negative predictive value
<sup>g</sup> Fisher's Exact test for independence was used, given rare exposure and rare outcome. This tested the null hypothesis that there was no difference in SARS-CoV-2 infection for people who would have been prohibited entry to campus as a result of screening for fever using the strategy in question, and those who would be permitted entry to campus.

There was an association between temperature readings and SARS-CoV-2 PCR result, with an increase of 0.1°F above individual mean body temperature the day of the PCR specimen collection associated with a 1.11 increased odds of SARS-CoV-2 positivity (95% CI 1.06 – 1.17), which did not change when controlling for local ambient temperature and age.

## INTERPRETATION

We found that daily temperature monitoring to screen for SARS-CoV-2 infection – either to prompt symptom-based testing for diagnosis and contact tracing or to prevent spread through encouraging someone to remain in isolation – was feasible and acceptable to a wide variety of people affiliated with a large public university. However, almost 3 out of 4 people, particularly students, did not already own thermometers, and purchasing and disseminating thermometers of sufficient quality during a global pandemic proved nearly impossible: taken together, these thermometers cost over *18,200 just to cover the needs of our study participants, calling into question the feasibility of this strategy on a campus-wide level. This highlights the challenges of accessing this type of equipment during a true public health emergency (particularly a global one), and underscores one of the challenges to requiring self-monitoring of body temperature as a condition of entry to a college campus, i.e. it is likely unrealistic to expect accurate responses to required self-attestations of being fever and symptom-free, especially for students.

Overall, the sensitivity of temperature or symptoms screening in our study never rose above 40.5%, and the best-performing quantitative temperature threshold had a sensitivity of only 30.6%, but considered fever to be anything at or above 98.7°F, which would be considered normal body temperature in most settings and resulted in a positive predictive value of only 3.5% (95% CI 2.2 – 5.8%). These findings align well with recently published results from an Australian hospital, where fever ≥100.4°F was only detected in 16 of 86 patients testing positive for SARS-CoV-2 over a 2-month period (sensitivity of 19%, 95% CI 11-28%), when using a variety of in-hospital temperature measurement methods, mostly with temporal thermometers.^37^ While their sensitivity using this fever threshold was higher than we observed in this study, this is unsurprising given that these were among hospital patients, not people who were mostly feeling well at time of testing. While the specificity of the strategies tested in our study was reasonably good (>93% in all cases), this study was conducted during the summer months, well before the typical influenza season in the San Francisco Bay Area. Specificity of temperature monitoring for SARS-CoV-2 can be expected to worsen in the fall and winter months, when the prevalence of influenza-like illness for reasons other than infection with SARS-CoV-2 is substantially increased.

There were significant associations between measured body temperature and PCR test results in longitudinal mixed models when controlling for individual variation in body temperatures; however, these regression models were designed to account for within-person variation in baseline temperature that is not scalable to real-life monitoring strategies, and does not apply to the most common temperature monitoring policies, which use dichotomous fever thresholds for permitting building entry or triggering symptoms-based SARS-CoV-2 testing, rather than a more sophisticated and individualized algorithm. Further, the performance of self-reported qualitative fever symptoms (“feeling feverish”) in this study suggests that daily quantitative temperature monitoring may not offer additional protection as a screening measure to prevent SARS-CoV-2 transmission on a college campus. Research by others that has found substantial measurement error with regard to body temperature^26^ bolsters the idea that qualitative self-report may be a preferable strategy.

Our study had a number of limitations. First, temperature data included here could be inaccurate for multiple reasons, including error using the thermometer or reporting error, such as participants not recording their temperature despite taking it or recording the temperature incorrectly. However, such limitations also apply to any scaled, non-research campus-level system for temperature monitoring. Second, while participants were asked to take their temperature in the morning, we did not control for time of day in this analysis, because there was substantial information bias in the daily survey timestamp (i.e. some participants anecdotally reported taking their temperature in the morning as requested but completing the survey at a later time). This may have made us less likely to detect fevers among our participants;^24^ however, since many school or work-based temperature screenings likely take place in the morning, this limitation would also apply to any scaled campus-level temperature screening system. Similarly, we did not control for differences in thermometer type in our analysis, given that the large majority of participants had an oral thermometer, despite the fact that type of thermometer used is related to mean body temperature measurements.^38^ However, this too is a limitation that would apply in real-life application of any campus-level temperature monitoring strategy. Third, individual basal temperature is known to vary by fertility cycle for women not taking hormones;^39^ however, we did not collect information about birth control or other hormone use during this study, so could not account for this in our analysis. A sensitivity analysis (not shown) stratifying our adjusted regression model by sex showed nearly identical results for women compared to the overall study population, suggesting that this hormonal temperature variation was not an important factor. Fourth, our results related to feasibility and acceptability of temperature monitoring were likely biased by the incentives provided to participants to encourage daily monitoring, as well as the enthusiasm for such activities among the type of person who would enroll in an intensive study of this nature, compared to a general campus population. However, the motivation to protect oneself during a pandemic and generally contribute to wellness of fellow members of the campus community can also reasonably be expected to make temperature monitoring more acceptable in the general population than might otherwise be true. Finally, it is possible that some participants had SARS-CoV-2 infection that was not detected by PCR testing, and therefore our effectiveness results may be biased in an unpredictable direction.

## Conclusions

In concordance with evidence from prior global pandemics,^6,8-11^ our findings suggest temperature screening using dichotomous fever thresholds during the COVID-19 pandemic may be little more than “security theater.”^40^ This term, largely attributed to computer security expert Bruce Schneier, describes measures that provide a *sense* of security, despite having no actual positive impact on security. Temperature monitoring strategies may be a rational response to community calls for action, even if those demands for action are based on inaccurate or outdated information about the risks and effective strategies for COVID-19 mitigation.^41,42^ Millions of dollars are invested in similar “security theater” measures each year in the United States, from radio-frequency identification (RFID) ankle bracelets to prevent hospital newborn abductions to removing shoes and prohibiting liquids above 3.4 ounces at airport security,^43,44^ in order to reassure the public that sufficient action is being taken in the face of a serious threat. Measures like this can have some benefit; namely, some individuals with SARS-CoV-2 would undoubtedly be detected by temperature screening measures and prevented from transmitting the virus to others. Further, routine temperature monitoring can reassure members of a campus community or workplace that those in charge care about their health and are taking the pandemic seriously. Yet an ineffective action can be harmful, if people believe that because temperature screening measures are in effect they are safe, and therefore other measures to prevent spread of SARS-CoV-2 – such as wearing a face covering and practicing social distancing – are not taken seriously.^45^ Further attention is needed to the benefits and drawbacks of various strategies to detect and prevent transmission of SARS-CoV-2 on college campuses and in similar close-knit communities, to ensure the community is fully aware of the practical and behavioral implications of any strategies employed. Colleges may want to consider using qualitative measures for self-report of feverishness as a symptom rather than attempting quantitative temperature monitoring, and focus resources on additional strategies known to prevent SARS-CoV-2 transmission, such as low-barrier access to SARS-CoV-2 testing without requiring disclosure of specific risks or symptoms,^46^ rather than relying on temperature screening or self-attestations of good health as a condition of campus access.

## Data Availability

De-identified data collected for this study, including individual participant data and a data dictionary defining each field in the set, will be made available to other researchers upon request. These data will be available beginning July 1, 2021 or upon publication of the final manuscript planned by key study staff, whichever comes first. Data can be accessed via email to the corresponding author, with a signed data access agreement and approval of a proposal by the study's Principal Investigators.

## ACKNOWLEDGMENTS

This work could not have been done without our hard-working graduate student researchers, who worked all summer at the specimen collection site and responded to participant questions and concerns about thermometers: Mariah De Zuzuarregui, Darren Frank, Sarah Gomez, Ariel Muñoz, Ruben Prado, Lawrence Tello, Emily Wang, and Sabrina Williamson. We also wish to thank our collaborators at UC Berkeley’s University Health Services, where specimen collection and thermometer pickup took place, including but not limited to: Judith Sansone, Melody Heller, Holly Stern, Tyler Crooks, Desi Gallardo, Jeff Kreutzen, Rebecca Stephenson, Lisa Polley, and Melissa Hennings; and our collaborators at the Innovative Genomics Institute Testing Consortium, including Fyodor D. Urnov, Jennifer A. Doudna, Alexandra M. Amen, Kerrie W. Barry, John M. Boyle, Cara E. Brook, Seunga Choo, L.T. Cornmesser, David J. Dilworth, Alexander J. Ehrenberg, Indro Fedrigo, Skyler E. Friedline, Thomas G.W. Graham, Ralph Green, Jennifer R. Hamilton, Ariana Hirsh, Megan L. Hochstrasser, Dirk Hockemeyer, Netravathi Krishnappa, Azra Lari, Hanqin Li, Enrique Lin-Shiao, Tianlin Lu, Elijah F. Lyons, Kevin G. Mark, Lisa Argento Martell, A. Raquel O. Martins, Patrick S. Mitchell, Erica A. Moehle, Christine Naca, Divya Nandakumar, Elizabeth O’Brien, Derek J. Pappas, Kathleen Pestal, Diana L. Quach, Benjamin E. Rubin, Rohan Sachdeva, Elizabeth C. Stahl, Abdullah Muhammad Syed, I-Li Tan, Amy L. Tollner, Connor A. Tsuchida, C. Kimberly Tsui, Timothy K. Turkalo, M. Bryan Warf, Oscar N. Whitney, and Lea B. Witkowsky. Last but not least, thanks to the specialists in Supply Chain Management at UC Berkeley who managed to find thousands of low-cost thermometers during a pandemic, particularly Stacey Templeman.

